# Soil-transmitted helminth reinfection after anthelmintic treatment in schools with improved versus unimproved WASH infrastructure in Mozambique: a prospective cohort study

**DOI:** 10.64898/2026.09.02.26362032

**Authors:** Valdemiro Novela, Augusto Messa, Pedro Emmanuel Fleitas, Berta Grau-Pujol, Javier Gandasegui, Charfudin Sacoor, Anélsio Cossa, Osvaldo Muchisse, José Carlos Jamine, Maria Martinez-Valladares, Khátia Munguambe, Inácio Mandomando, Stephen R. Doyle, Jose Muñoz

## Abstract

**Background:** Soil-transmitted helminths (STH) infect more than a billion people worldwide, disproportionately affecting communities with poor water, sanitation, and hygiene (WASH). Mass drug administration is the primary control strategy, but reinfection frequently occurs within months of treatment, and the extent to which improved WASH reduces reinfection risk remains unclear. We conducted a prospective cohort study comparing STH reinfection rates over seven months after anthelmintic treatment among school-aged children attending schools with improved versus unimproved WASH in the Manhiça district, Mozambique, using reinfection dynamics as a proxy for the impact of WASH interventions.

**Methods:** Children were screened for STH using Kato-Katz microscopy, then dewormed with albendazole and praziquantel (plus ivermectin for those positive for *T. trichiura* and/or *S. stercoralis*). A follow-up sample was collected 21 days post-treatment from initially positive children to assess the cure rate (CR) and egg reduction rate (ERR). Cured participants were enrolled in the WASH-improved or WASH-unimproved cohort and followed monthly for seven months. Reinfection and incidence rates were estimated, and Cox proportional hazards regression was used to assess the association between WASH cohort and time to reinfection.

**Results:** STH prevalence was 17.5% (95% CI: 15.2–19.8; *n*=178/1018), comprising *T. trichiura* (9.5%), *A. lumbricoides* (6.3%), *S. stercoralis* (2.6%), and hookworms (1.9%). No heavy-intensity infections were observed; most *T. trichiura* (94.9%) and hookworm (94.7%) infections were light, whereas 43.8% of *A. lumbricoides* infections were moderate. CR/ERR were 88.4%/96.56% for *T. trichiura* (n=95) and 93.3%/93.36% for *A. lumbricoides* (n=50). Overall reinfection rate was 32.3% (n=54/167), higher in the improved-WASH cohort (39.3%) than in the unimproved-WASH cohort (18.2%), corresponding to 66.38 reinfections per 1,000 person-months at risk overall (82.27 vs. 35.89 by cohort). The unimproved-WASH cohort had a lower hazard of reinfection (HR=0.456; 95% CI: 0.229–0.908; *p*=0.0254).

**Conclusions:** Despite effective treatment, over a third of children were reinfected within seven months, with reinfection unexpectedly higher in the improved-WASH cohort. This suggests that infrastructure-based WASH classification may not capture true exposure or behavioural risk, underscoring the need for household-level monitoring to assess WASH impact alongside MDA. Clinical trial number: not applicable.

## Background

Soil-transmitted helminths (STH) are a group of gastrointestinal nematodes comprising the whipworm *Trichuris trichiura*, the roundworm *Ascaris lumbricoides*, the two hookworms *Necator americanus* and *Ancylostoma duodenale*, and the threadworm *Strongyloides stercoralis*. Collectively, STH are the most prevalent of the Neglected Tropical Diseases (NTDs) (1,2). The 2021 Global Burden of Disease study estimated that STH infect around 643 million people worldwide (3). Infections with *S. stercoralis*, often overlooked among the STHs and considered the neglected of the NTDs, are estimated to affect an additional 386 million people (1,4,5). Signs and symptoms of infection vary by STH species and infection intensity; however, morbidity includes gastrointestinal distress, nutritional deficiencies and anaemia, malnutrition, impaired growth, and impaired cognitive development, particularly in children (1,6). In high-burden infections and in the presence of comorbidities, morbidity can lead to more serious disease, including in adults.

In the 2021-2030 World Health Organization (WHO) road map for NTDs, STHs are targeted for elimination as a public health problem by 2030, defined as <2% of moderate-to-heavy intensity infections (7). This is to be achieved through preventive chemotherapy (PC) with the anthelmintic drugs albendazole or mebendazole in areas where prevalence exceeds 20%, delivered through annual or biannual mass-drug administration (MDA) campaigns targeting pre-school (pre-SAC) and school-aged children (SAC), women of childbearing age, and adults in certain high-risk occupations (1,8). This approach has been effective in controlling morbidity, but reinfection is likely to persist if people continue to live in highly contaminated environments due to inadequate WASH infrastructure. Therefore, interventions involving health education and WASH are also necessary, underscoring the limitations of traditional control measures and the urgent need to develop and implement more effective, innovative strategies (9,10). Accordingly, the WHO also set the target that by 2030, access to at least basic sanitation and hygiene in regions endemic for STH is universal (7).

In 2024, it was estimated that 74% of the global population had access to safely managed drinking water, 58% used safely managed sanitation services, and 80% had access to basic hygiene facilities. Coverage is substantially lower in Sub-Saharan Africa (32%, 26%, and 27%, respectively) and lower still in Mozambique for water and hygiene (28% and 16%), though sanitation coverage (34%) exceeds the regional average (11). WASH is thought to reduce STH transmission through two complementary mechanisms: sanitation limits environmental contamination by infected individuals, while hygiene reduces individuals’ exposure to that contamination; water access underpins both (12). However, evidence for this relationship is mixed depending on study design: meta-analyses have shown a positive impact on reducing prevalence and intensity of infection (13,14), and modelling suggests that WASH will be pivotal in the long-term to sustain control or elimination of infection after preventive chemotherapy is scaled down (12), whereas large randomized controlled trials and longitudinal studies have not supported this association between WASH improvement and STH reduction (15,16). Given this inconsistency, it is critical to understand the extent to which WASH contributes to STH transmission in low-income countries such as Mozambique, where this question has remained unexplored to date.

Herein, we describe a prospective cohort study conducted in the Manhiça district, Southern Mozambique, in which we measured anthelmintic treatment efficacy and STH reinfection rates over seven months among school-aged children attending schools with contrasting WASH conditions, using reinfection as a proxy to assess the extent to which WASH improvements reduce STH transmission.

## Methods

### WASH-it study design and setting

We conducted a prospective cohort study, WASH-it, in the Manhiça district, approximately 80 km north of the capital, Maputo, in southern Mozambique, between February 2019 and February 2020. The district covers approximately 2,380 km2 and has a subtropical climate with two distinct seasons: a warm, rainy season from November to April, and a cool, dry season for the rest of the year (17). Since 1996, the Centro de Investigação em Saúde de Manhiça (CISM) has managed a Health and Demographic Surveillance System (HDSS) in the Manhiça district, which follows 201,845 individuals in 46,441 enumerated, geo-positioned households, from which regular updates on all demographic events are collected (18). A previous cross-sectional study in this area reported prevalence of at least one STH of 13.1% (overall), 11.5% (children 5-15 years old), and 14.6% (over 15 years old) using the Kato-Katz method on a single stool (19).

The WASH-it study aimed to evaluate the association between improvements in water, sanitation, and hygiene conditions in schools and reinfection with *T. trichiura*, *A. lumbricoides*, hookworms, and *S. stercoralis* in schoolchildren during the seven months following anthelmintic treatment, as a proxy for the impact of a WASH intervention. To achieve this, children from two schools, within CISM’s HDSS, but separated by 57.5 km, namely the “Escola Primária Completa de Pateque” in the Maluana administrative post and the “Escola Primária Completa de Taninga” in the 3 de Fevereiro administrative post, were included. Both schools – henceforth referred to as Pateque and Taninga, respectively – hosted a similar number of pupils (1,015 and 908, respectively). Before study initiation, a WASH improvement intervention was conducted by a non-governmental organization (NGO) in Pateque and the surrounding community, between November 2017 and November 2018. The intervention consisted in (i) providing a clean water supply, (ii) constructing improved latrines, and maintaining a minimum ratio of latrines per child as recommended by UNICEF (1 latrine per 25 children), (iii) promoting health and maintenance of the latrines, including the organization of a group of volunteers to keep the latrines clean following the “Community-Led Total Sanitation (CLTS)” method (20), (iv) constructing five new deep wells for improved water supply in the community, (v) covering five uncovered deep wells in the community, and (vi) promoting the construction and maintenance of improved latrines in the community. Taninga, on the other hand, did not have access to these improved WASH conditions.

Between 13 February and 09 July 2019, children registered at these schools were screened for STH by microscopy, as detailed below. All children were dewormed, and a follow-up sample was collected from those who tested positive at screening to assess treatment response. If infection was cleared (cured), participants were enrolled in the WASH-improved or WASH-unimproved cohorts for follow-up over the next seven months, with monthly stool samples collected to determine STH reinfection.

### Study population and sample size justification

The study was conducted among school-aged children enrolled in the two schools. To be included, children had to be (i) enrolled in CISM’s HDSS, (ii) aged between 6 and 18 years (both ages included), (iii) from the schools under study, and (iv) have parental consent to participate in the study, as evidenced by written informed consent. Participants who indicated an intention to move to another household, neighbourhood, or school within the next 6 months were excluded from the study.

We aimed to recruit 100 STH-infected participants per cohort (improved and unimproved WASH). This sample size would provide 80% power to detect a minimum difference of 18% in the reinfection rate between the null hypothesis (the reinfection rate in both cohorts is 40%) and the alternative hypothesis (the reinfection rate in the improved WASH cohort is 22% or lower), using a two-sided Chi-square test at a significance level of 0.05. To recruit 100 participants per cohort, approximately 1,100 children (550 in each school) were enrolled, based on an estimated STH prevalence of 20% in these populations and allowing for a 10% loss to follow-up.

### Study procedures

Before study activities began, schools were visited to explain the study objectives and approach to teachers and other members of the school community, and to seek permission at the community and school levels. A questionnaire containing school information was also completed. As children attending the schools (improved and unimproved) were under 18 years old, parents and guardians were invited to attend a meeting at the school, where the study was explained and informed consent was obtained. A study questionnaire for each child was then completed. A sample collection kit (a sterile flask for stool and two pairs of gloves for protection) was provided to each child-parent/guardian pair, and instructions were given on how to collect stool samples. The following day, a field worker returned to the school, collected stool samples from each child, and transported them to the laboratory in a cooler box within four hours of collection.

STH infections were assessed by microscopy using the Kato-Katz and Baermann techniques (see below for details). Two stool aliquots, without preservatives, were stored at –80 °C for subsequent use. After the baseline assessment, all children in both schools were dewormed with a single dose of albendazole (400 mg) to treat *A. lumbricoides* and hookworm infections, and a single dose of praziquantel (40 mg/kg) for *Schistosoma* spp. infections, as per national guidelines. For children positive for *T. trichiura* or *S. stercoralis*, an additional dose of ivermectin (200 µg/kg) was provided. Twenty-one days after treatment, participants who were STH-positive at baseline were asked to provide a second stool sample to assess treatment response. Those who tested negative after treatment and were thus considered cured were included in the “reinfection” follow-up cohorts (improved WASH and unimproved WASH); those who remained positive were treated again with albendazole and ivermectin, if necessary (i.e. positive for *T. trichiura* and/or *S. stercoralis*). A new sample was collected and assessed by microscopy, and participants were included in the cohorts if negative after the second treatment. During the seven months after treatment, a single stool sample was collected every 30 days to assess for STH reinfection using the same collection and microscopic examination methods. Once cohort participants were found to be reinfected, they were treated with albendazole, according to local STH management protocols, and follow-up was stopped. A summary of study procedures is provided in the flowchart in Figure 1.

**Figure 1.**
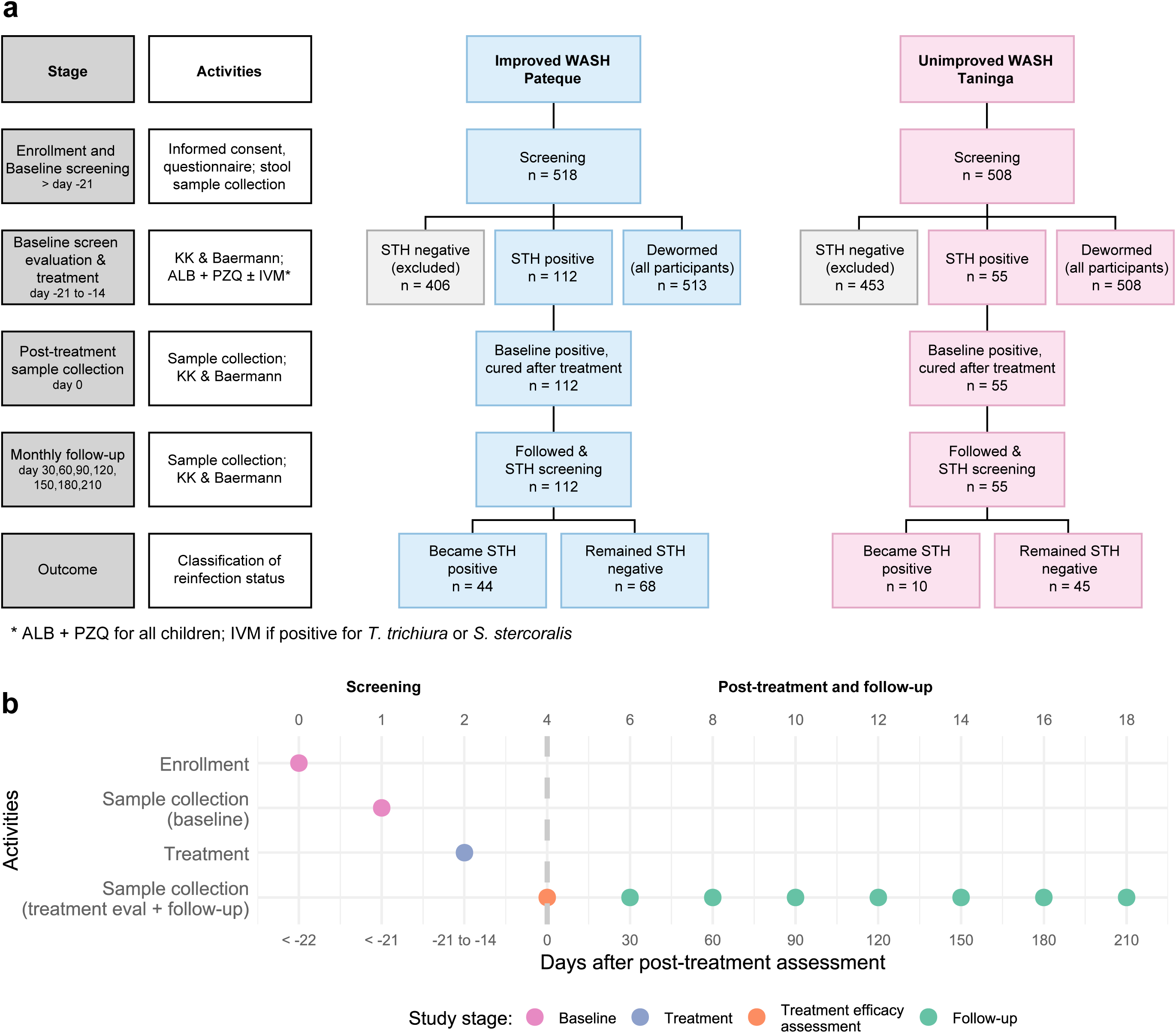
Diagram of the recruitment flow for 6-to 18-year-old participants for the WASH-it study. **a**) Study activities flow for the two cohorts (Pateque and Taninga). b) Timeline of study activities per individual participants. *Treatment was provided with albendazole (ALB) and praziquantel (PZQ) as per national MDA guidelines. Ivermectin was provided to participants who tested positive for *T. trichiura* and *S. stercoralis*. KK-Kato-Katz. Five children enrolled in Pateque could not be located after screening, because they either had quit attending school or had been transferred to a different school.

#### Laboratory methods

Stool samples were examined using duplicate Kato-Katz thick smears with the 41.8 mg template (Sterlitech Corporation, USA) to detect and quantify eggs of *T. trichiura*, *A. lumbricoides*, and hookworms. To assess infection intensity, egg counts per slide from Kato-Katz thick smears were multiplied by 24 to obtain the number of eggs per gram of faeces (21). Additionally, the Baermann method was used to detect *S. stercoralis* larvae in stool samples. This was performed by mixing 10 g of the stool sample with 2 g of activated charcoal and lukewarm water, then transferring the mixture to a Petri dish lined with a double layer of tissue paper (bottom) and covered with a single layer of paper at the top to form a small pouch. After an 18-24-hour incubation at 26 °C, the mixtures were suspended for 1 hour in lukewarm water at room temperature and filtered using a conventional Baermann apparatus (a strainer on top of a funnel connected to a rubber hose clamped with a haemostatic clamp) supported by a funnel stand. The lower 10 mL of water in the hose was drained off, centrifuged at 2000 rpm for 5 minutes, and the remaining ∼1 mL of the sediment was examined microscopically for the presence of larvae (22). *S. stercoralis* larvae were identified by adding a drop of Lugol and observing under the microscope to assess morphological characteristics (22). For quality control, a second microscopist was required to observe the larvae and confirm the identification.

### Data analysis

Univariate analysis was conducted to describe the participant population, with 95% confidence intervals (CI) reported where relevant. Frequencies were compared using two-sided Chi-square or Fisher’s exact tests, and p-values were adjusted for multiple comparisons using the Holm-Bonferroni method where applicable. Kato-Katz egg count data were summarised by species and by group (sex and enrolment school) using arithmetic and geometric means. Because some participants had zero eggs in the post-treatment stool sample, a value of 1 was added to egg counts to enable logarithmic transformation.

WHO thresholds for classifying infection intensity (light, moderate, or heavy) are based on Kato-Katz faecal egg counts, expressed as eggs per gram (EPG) of faeces. These thresholds are defined by species as follows: *T. trichiura*: light (1-999 EPG), moderate (1,000-9,999 EPG) and heavy (≥10,000 EPG); *A. lumbricoides*: light (1-4,999 EPG), moderate (5,000-49,999 EPG) and heavy (≥50,000 EPG); and Hookworm: light (1-1,999 EPG), moderate (2,000-3,999 EPG) and heavy (≥4,000 EPG) (23).

Treatment response was assessed using the cure rate (CR), defined as the percentage of participants cured, i.e., those with no eggs observed in the Kato-Katz thick smear or Baermann for the given species in the post-treatment sample (21 days after treatment) (24).

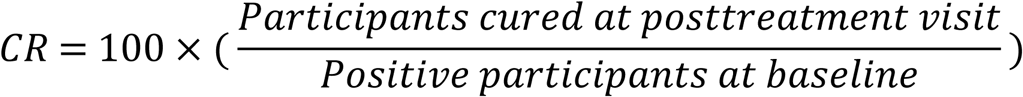

The egg reduction rate (ERR) was calculated to assess the decrease in the egg burden of *T. trichiura*, *A. lumbricoides*, and hookworms following treatment, using the arithmetic and geometric means, with the formula (23):

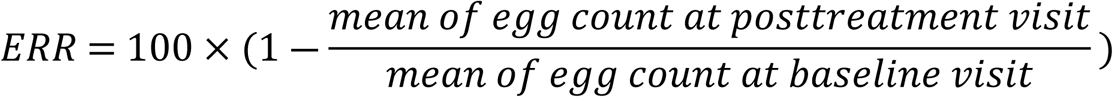

Reinfection rates were calculated for participants cured after treatment with anthelmintic drugs and followed monthly until reinfection or the last study visit (seven months post-treatment). Additionally, reinfection incidence rates (IR) were estimated based on the duration each participant remained negative after infection clearance following treatment (i.e., no observable eggs or larvae on Kato-Katz or Baermann in the post-treatment sample). This time contribution was measured until they were no longer followed, became positive, or the last study visit (seven months post-treatment), whichever occurred first. We expressed reinfection incidence rates in 1,000 person-months at risk (PMAR), using the following formula:

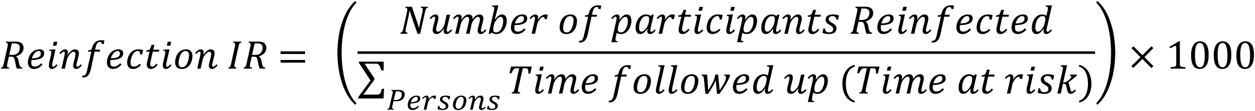

Cumulative reinfection rates were estimated using Kaplan-Meier curves. Cox proportional hazards regression was used to assess whether enrolment in improved or unimproved WASH was associated with time to reinfection with STH (all species combined and separately). Participants who were positive, successfully treated, and did not provide additional stool samples were considered lost to follow-up and excluded from the reinfection analysis. All statistical analyses were performed in R (version 4.5.0) (25).

## Results

### Participant characteristics

During screening, 1,038 participants were invited to take part in the study. Of these, 1,026 were enrolled at the baseline visit, with 518 in Pateque (improved WASH) and 508 in Taninga (unimproved WASH). Nearly 54% (*n*=553) of enrolled participants were female, and the mean age was 9.5 years (SD=2.56). Around half of participants were aged 6-9 years (52%; *n*=534), followed by those aged 10-13 years (41.5%; *n*=426), with those aged 14-18 years the least frequently enrolled (6.4%; *n*=66). These characteristics were similar across participants enrolled in each school. Most pupils reported handwashing before meals and after urination/defecation, while fewer reported handwashing on waking (0.9%), before dinner (56.6%), or before sleeping (3.3%). Overall, 42.9% reported defecating in a latrine (improved or unimproved) at school; <2% reported open defecation (inside or outside the school yard). Most pupils wore footwear and presented with a clean face during the interview, with only a few reporting diarrhoea or pruritus. The population characteristics of study participants are presented in Table 1 and in Table 1 of Additional File 1.

**Table 1.** Baseline characteristics of study participants for the WASH-it study.

| Variable | Category | Pateque (improved WASH) | Tanninga (unimproved WASH) | Overall |
| --- | --- | --- | --- | --- |
| Subjects | N | 518 | 508 | 1026 |
| Sex | Female | 54.8% (284) | 53% (269) | 53.9% (553) |
|  | Male | 45.2% (234) | 47% (239) | 46.1% (473) |
| Age in years | Mean ± SD | 9.5 ± 2.6 | 9.5 ± 2.51 | 9.5 ± 2.56 |
| Age group | 6-9 years | 51.7% (268) | 52.4% (266) | 52% (534) |
|  | 10-13 years | 40.9% (212) | 42.1% (214) | 41.5% (426) |
|  | 14-17 years | 7.3% (38) | 5.5% (28) | 6.4% (66) |

### Prevalence and burden of individual STH infections

The overall prevalence of infection with at least one STH was 17.5% (95% CI: 15.2-19.8; *n*=178/1,018). *T. trichiura* (9.5%, 95% CI: 7.7-11.3; *n*=97/1,021) was the most frequent species, followed by *A. lumbricoides* (6.3%, 95% CI: 4.8-7.8; *n*=64/1,021), *S. stercoralis* (2.6%, 95% CI: 1.7-3.6; *n*=27/1,023), and hookworms (1.9%, 95% CI: 1-2.7; *n*=19/1,021) as the least frequent (Table 2). The prevalence of STH was significantly higher in the school with improved WASH (23.3%, 95% CI: 19.7-27; *n*=119/510) than in the one with unimproved WASH (11.6%, 95% CI: 8.8-14.4; *n*=59/508) (*p*=7.25×10-7). This pattern also held for *T. trichiura* and *A. lumbricoides*, but the prevalence of *S. stercoralis* was higher in the school with unimproved WASH than in the one with improved WASH (*p*=7.31×10^-4^). No significant differences were observed for hookworms.

**Table 2.**
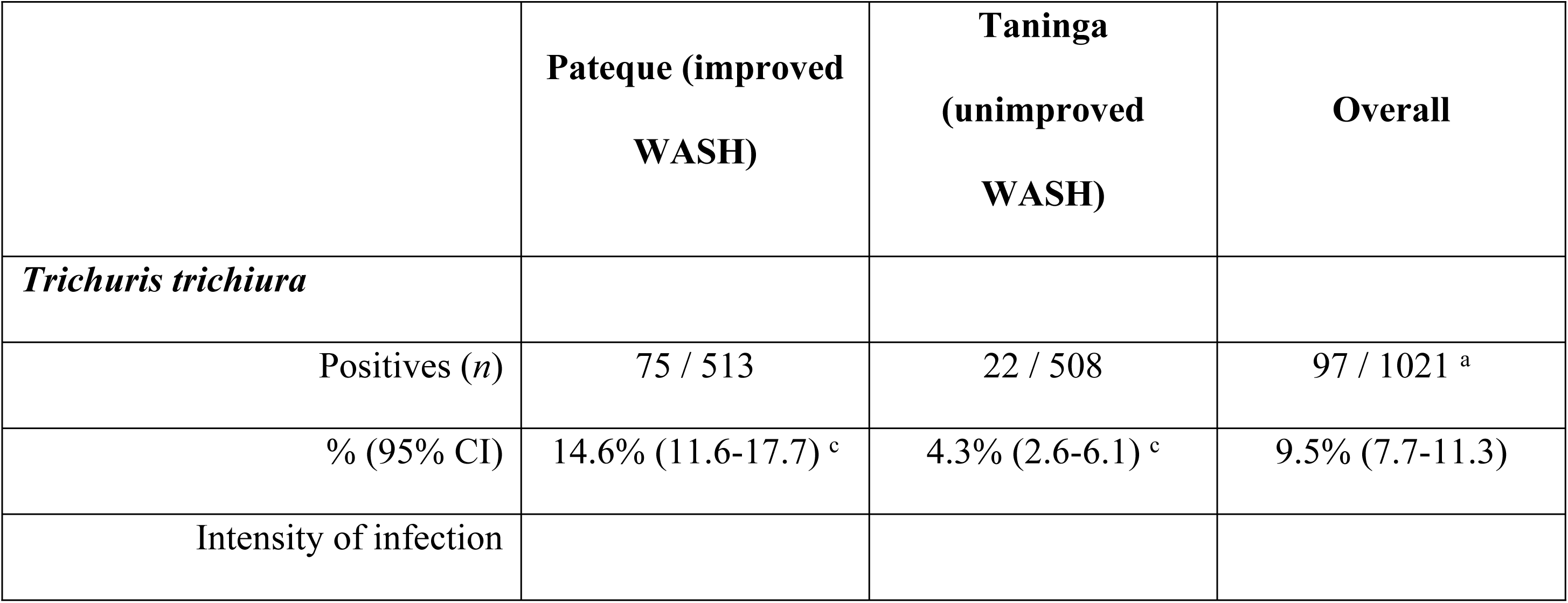
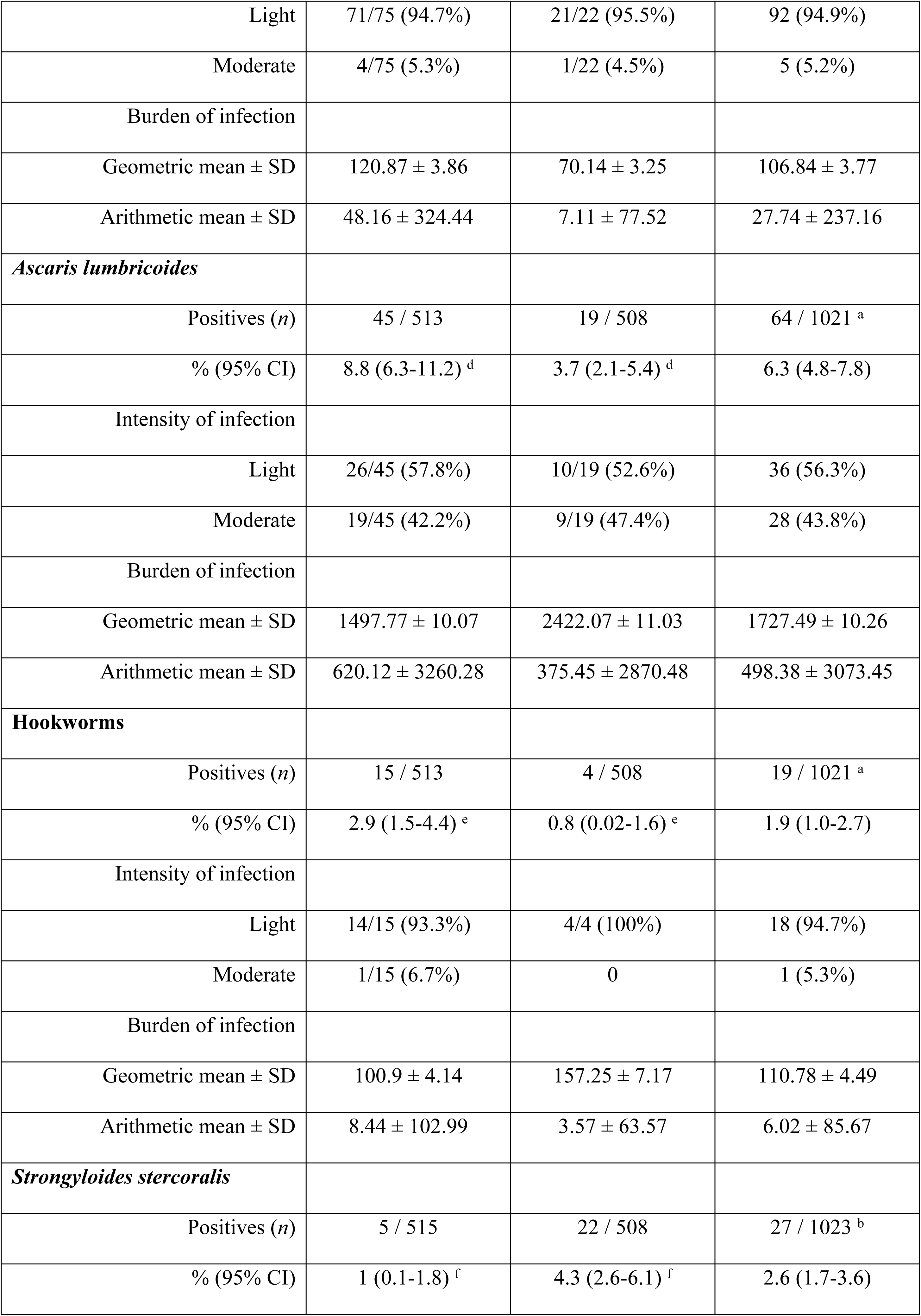
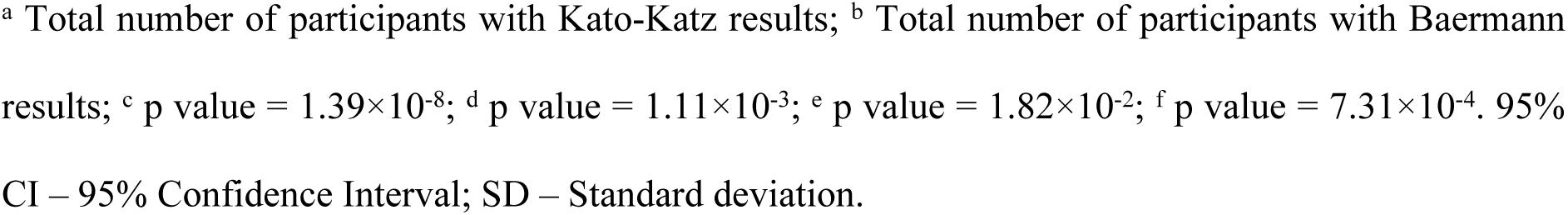
Prevalence, intensity, and burden of STH infection at baseline.

|  | Pateque (improved WASH) | Tanninga (unimproved WASH) | Overall |
| --- | --- | --- | --- |
| *Trichuris trichiura* |  |  |  |
| Positives (n) | 75 / 513 | 22 / 508 | 97 / 1021 <sup>a</sup> |
| % (95% CI) | 14.6% (11.6-17.7) <sup>c</sup> | 4.3% (2.6-6.1) <sup>c</sup> | 9.5% (7.7-11.3) |
| Intensity of infection |  |  |  |
| Light | 71/75 (94.7%) | 21/22 (95.5%) | 92 (94.9%) |
| Moderate | 4/75 (5.3%) | 1/22 (4.5%) | 5 (5.2%) |
| Burden of infection |  |  |  |
| Geometric mean ± SD | 120.87 ± 3.86 | 70.14 ± 3.25 | 106.84 ± 3.77 |
| Arithmetic mean ± SD | 48.16 ± 324.44 | 7.11 ± 77.52 | 27.74 ± 237.16 |
| ***Ascaris lumbricoides*** |  |  |  |
| Positives (*n*) | 45 / 513 | 19 / 508 | 64 / 1021 <sup>a</sup> |
| % (95% CI) | 8.8 (6.3-11.2) <sup>d</sup> | 3.7 (2.1-5.4) <sup>d</sup> | 6.3 (4.8-7.8) |
| Intensity of infection |  |  |  |
| Light | 26/45 (57.8%) | 10/19 (52.6%) | 36 (56.3%) |
| Moderate | 19/45 (42.2%) | 9/19 (47.4%) | 28 (43.8%) |
| Burden of infection |  |  |  |
| Geometric mean ± SD | 1497.77 ± 10.07 | 2422.07 ± 11.03 | 1727.49 ± 10.26 |
| Arithmetic mean ± SD | 620.12 ± 3260.28 | 375.45 ± 2870.48 | 498.38 ± 3073.45 |
| **Hookworms** |  |  |  |
| Positives (*n*) | 15 / 513 | 4 / 508 | 19 / 1021 <sup>a</sup> |
| % (95% CI) | 2.9 (1.5-4.4) <sup>e</sup> | 0.8 (0.02-1.6) <sup>e</sup> | 1.9 (1.0-2.7) |
| Intensity of infection |  |  |  |
| Light | 14/15 (93.3%) | 4/4 (100%) | 18 (94.7%) |
| Moderate | 1/15 (6.7%) | 0 | 1 (5.3%) |
| Burden of infection |  |  |  |
| Geometric mean ± SD | 100.9 ± 4.14 | 157.25 ± 7.17 | 110.78 ± 4.49 |
| Arithmetic mean ± SD | 8.44 ± 102.99 | 3.57 ± 63.57 | 6.02 ± 85.67 |
| ***Strongyloides stercoralis*** |  |  |  |
| Positives (*n*) | 5 / 515 | 22 / 508 | 27 / 1023 <sup>b</sup> |
| % (95% CI) | 1 (0.1-1.8) <sup>f</sup> | 4.3 (2.6-6.1) <sup>f</sup> | 2.6 (1.7-3.6) |

<sup>a</sup> Total number of participants with Kato-Katz results; <sup>b</sup> Total number of participants with Baermann results; <sup>c</sup> p value = $1.39 \times 10^{-8}$ ; <sup>d</sup> p value = $1.11 \times 10^{-3}$ ; <sup>e</sup> p value = $1.82 \times 10^{-2}$ ; <sup>f</sup> p value = $7.31 \times 10^{-4}$ . 95% CI – 95% Confidence Interval; SD – Standard deviation.

No heavy-intensity infections were observed for any of the helminth species identified through Kato-Katz. Most *T. trichiura* (94.9%) and hookworm (94.7%) infections were classified as light-intensity, whereas for *A. lumbricoides*, 43.8% of infections were moderate-intensity. No significant differences in infection intensity were observed between schools of enrolment (Table 2). Egg counts per gram of stool ranged from 12-5,940 for *T. trichiura*, 12-39,192 for *A. lumbricoides*, and 12-2,148 for hookworms. Using arithmetic means, infection intensity was higher in children from the school with improved WASH who were infected with *T. trichiura, A. lumbricoides*, and hookworms. Using geometric means, infection intensity followed the same pattern for *T. trichiura* and hookworms; however, for *A. lumbricoides*, it was higher among children from the school with unimproved WASH.

Among the 179 participants infected with any STH, 84.9% harboured a single species, while co-infections with two and three species occurred in 14.5% and 0.01%, respectively (Figure 2). Among co-infected participants, the most common combination was *T. trichiura* and *A. lumbricoides* (n = 21), whereas all other STH combinations were observed in only one participant each (Figure 2). The sole participant co-infected with three species harboured *T. trichiura*, hookworms, and *S. stercoralis*. The distribution of co-infection combinations by school is shown in Table 2 (Additional File 1).

**Figure 2.**
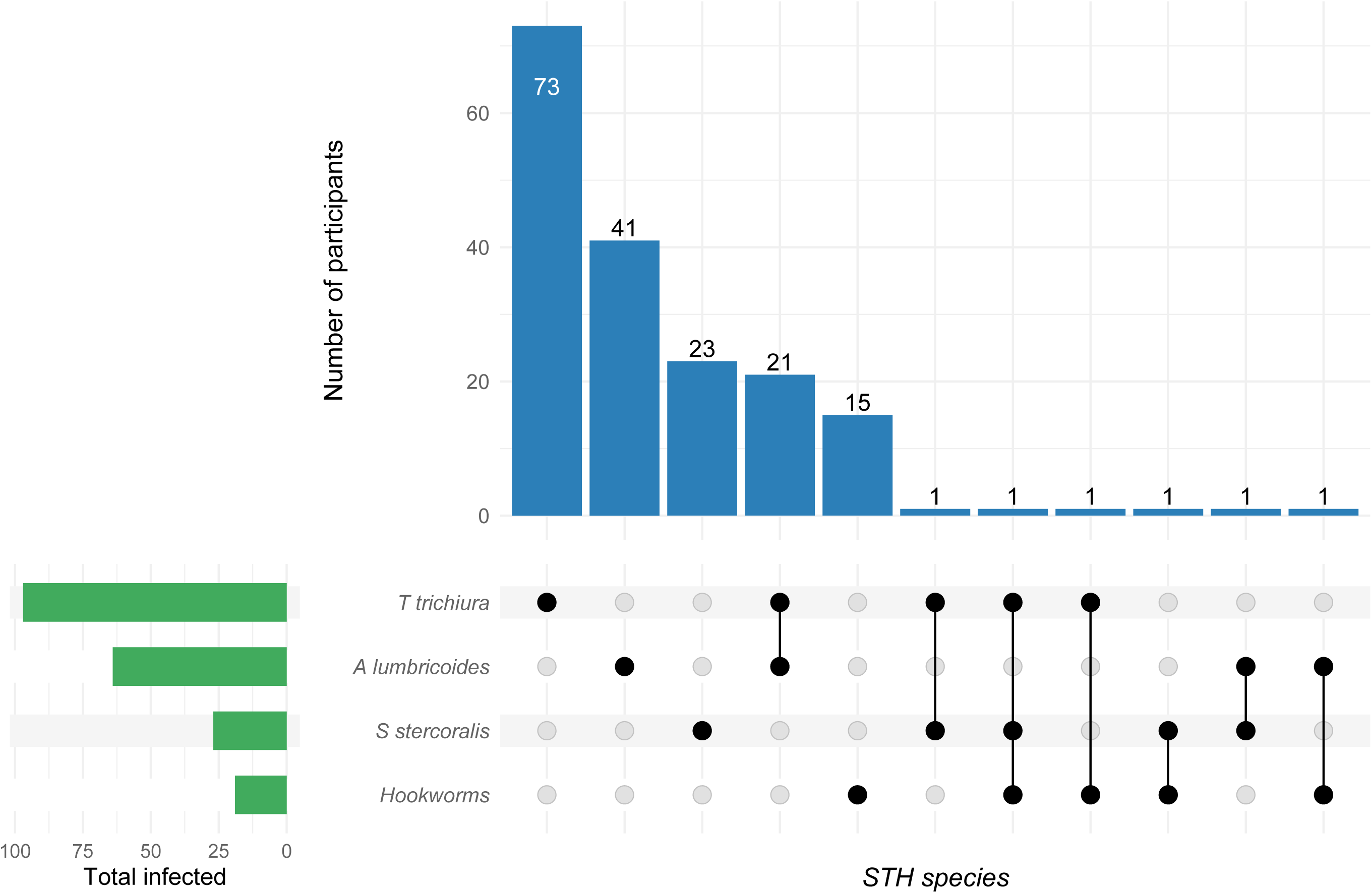
Frequency and distribution of co-infections among participants infected by at least one soil-transmitted helminth species (*n*=179) at the baseline assessment for the WASH-it study.

### Treatment response and reinfection by STH

Treatment response was assessed in participants with both baseline (screening) and post-treatment (Day 0) samples (Table 3). For participants infected with *T. trichiura* at baseline, treatment response could be assessed in 95/97 (97.9%) participants. Among these participants, the cure rate was 88.4% (95% CI: 80.2-94.1), whereas the egg reduction rates were 98.58% (95% CI: 97.97-99.00) and 96.56% (95% CI: 92.76-100) using geometric and arithmetic means, respectively. For 50/64 (78.1%) participants infected with *A. lumbricoides*, a CR of 93.3% (95% CI: 83.8-98.2) and ERRs of 99.9% (95% CI: 99.8-99.96) and 93.36% (95% CI: 84.35-100), using geometric and arithmetic means, respectively, were observed. For 19/19 (100%) participants infected with hookworms, the cure rate was 94.7% (95% CI: 73.9-99.9%), and the ERRs were 98.98% (95% CI: 97.74-99.55) and 99.8% (95% CI: 99.35-100), using geometric and arithmetic means, respectively. For *S. stercoralis*, 26/27 (96.3%) participants were assessed, and the cure rate was 96.2% (95% CI: 80.4-99.9).

**Table 3.** Cure rates (CR) and egg reduction rates (ERR) 21 days after anthelmintic treatment.

|  | **Pateque (improved**<br>**WASH)** | **Taninga**<br>**(unimproved**<br>**WASH)** | **Overall** |
| --- | --- | --- | --- |
| *Trichuris trichiura* |  |  |  |
| Number of participants positive |  |  |  |
| Before treatment | 73 | 22 | 95<sup>a</sup> |
| After treatment | 8 | 3 | 11 |
| Cure rate (95% CI) | 89% (79.5-95.1) | 86.4% (65.1-97.1) | 88.4% (80.2-94.1) |
| Geometric mean EPG |  |  |  |
| Before treatment | 125.08 | 71.83 | 107.97 |
| After treatment | 1.54 | 1.65 | 1.56 |
| Geometric mean ERR (95% CI) | 98.77% (98.15-99.18) | 97.70% (95.09-98.93) | 98.58% (97.97-99.00) |
| Arithmetic mean EPG |  |  |  |
| Before treatment | 333.5 | 164.18 | 294.32 |
| After treatment | 11.5 | 5.45 | 10.11 |
| Arithmetic mean ERR (95% CI) | 96.55% (92.21-100) | 96.68% (91.43-100) | 96.56% (92.76-100) |
| *Ascaris lumbricoides* |  |  |  |
| Number of participants positive |  |  |  |
| Before treatment | 41 | 19 | 60<sup>b</sup> |
| After treatment | 3 | 1 | 4 |
| Cure rate (95% CI) | 92.7% (80.1-98.5) | 94.7% (73.97-99.9) | 93.3% (83.8-98.2) |
| Geometric mean EPG |  |  |  |
| Before treatment | 1,608.14 | 2,434.72 | 1,822.40 |
| After treatment | 1.76 | 1.58 | 1.70 |
| Geometric mean ERR (95% CI) | 99.89% (99.74-99.95) | 99.94% (99.69-99.99) | 99.90% (99.80-99.96) |
| Arithmetic mean EPG |  |  |  |
| Before treatment | 7,662.73 | 10,038.32 | 8,415 |
| After treatment | 671.12 | 315.16 | 558.4 |
| Arithmetic mean ERR (95% CI) | 91.24% (77.42-100) | 96.86% (89.74-100) | 93.36% (84.35-100) |
| ***Hookworms*** |  |  |  |
| Number of participants positive |  |  |  |
| Before treatment | 15 | 4 | 19<sup>c</sup> |
| After treatment | 1 | 0 | 1 |
| Cure rate (95% CI) | 93.3% (68.1-99.8) | 100% (39.8-100) | 94.7% (73.9-99.9) |
| Geometric mean EPG |  |  |  |
| Before treatment | 102.89 | 160.89 | 110.78 |
| After treatment | 1.19 | 1 | 12 |
| Geometric mean ERR (95% CI) | 98.85% (97.14-99.53) | 99.37% (86.47-99.97) | 98.98% (97.74-99.55) |
| Arithmetic mean EPG |  |  |  |
| Before treatment | 288.8 | 453 | 323.37 |
| After treatment | 0.8 | 0 | 2.4 |
| Arithmetic mean ERR (95% CI) | 99.71% (99.03-100) | 100% (100-100) | 99.80% (99.35-100) |
| ***Strongyloides stercoralis*** |  |  |  |
| Number of participants positive |  |  |  |
| Before treatment | 4 | 22 | 26<sup>d</sup> |
| After treatment | 1 | 0 | 1 |
| Cure rate (95% CI) | 75% (19.4-99.4) | 100% (84.6-100) | 96.2% (80.4-99.9) |

<sup>a</sup> Three positive participants at baseline had no follow-up samples (0196, 0197, 0573) and were excluded; <sup>b</sup> Four positive participants at baseline had no follow-up samples (0196, 0279, 0445, 0573) and were excluded; <sup>c</sup> One positive participant at baseline had no follow-up sample (0197) and was excluded; <sup>d</sup> One positive participant at baseline had no follow-up sample (0101) and was excluded. EPG – eggs per gram of stool; ERR – Egg reduction rate.

Over the 7-month study period, the overall reinfection rate for STH was 32.3% (*n*=54/167), with a higher rate among children in the improved WASH cohort (39.3%; *n*=44/112) than among those in the unimproved WASH cohort (18.2%; *n*=10/55). By species, reinfection rates were 30.3% (*n*=27/89) for *T. trichiura*, 26.3% (*n*=15/57) for *A. lumbricoides*, 26.3% (*n*=5/19) for hookworms, and 12% (*n*=3/25) for *S. stercoralis* (Figure 3 and Table 4). Reinfection rates for *T. trichiura, A. lumbricoides*, and hookworms over time are presented in Table 3 and Figures 1-4 in Additional File 1. Overall, the reinfection incidence rate (IR) was 66.38 reinfections per 1,000 person-months at risk (PMAR); this was higher among children in the improved WASH cohort (82.27 reinfections per 1,000 PMAR) than among those in the unimproved WASH cohort (35.89 reinfections per 1,000 PMAR). Among the STH species, *T. trichiura* had the highest IR at 64.27 (95% CI: 40.83-87.73) reinfections per 1,000 PMAR, followed by *A. lumbricoides* at 55.75 (95% CI: 28.33-83.17) and hookworms at 53.76 (95% CI: 7.16-91.33) reinfections per 1,000 PMAR, while *S. stercoralis* had the lowest incidence at 24.08 (95% CI: – 2.84-50.99) reinfections per 1,000 PMAR (Table 4).

**Figure 3.**
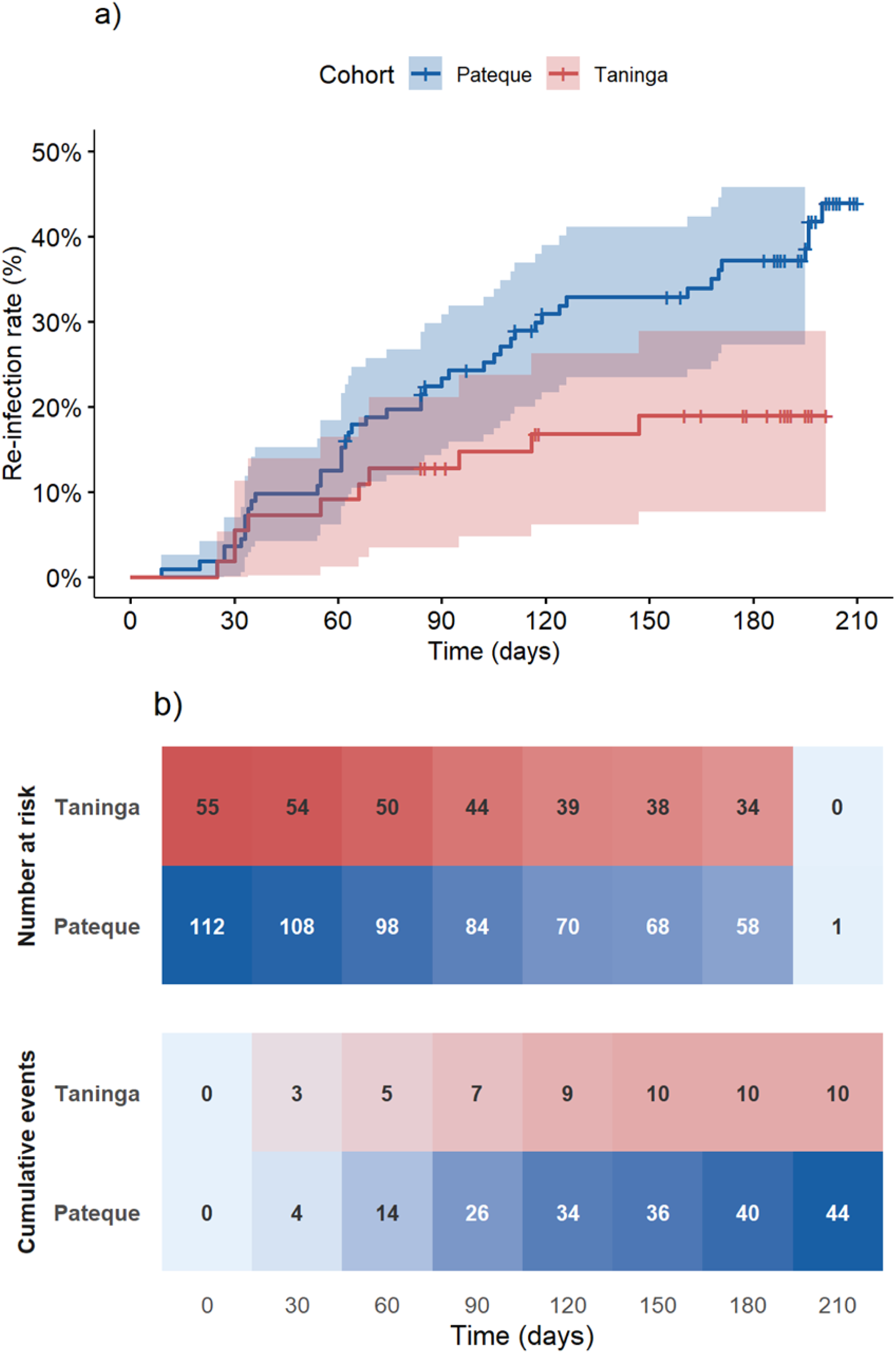
Reinfection rate by any soil-transmitted helminth in Pateque (improved WASH) and Taninga (unimproved WASH) cohorts throughout the seven-month follow-up period. **a**) Reinfection rates by any STH species in improved WASH cohort vs. unimproved WASH cohort; b) Progression of number of participants at risk and cumulative or reinfection events for participants in Pateque and Taninga.

**Table 4.** STH reinfection incidence rates.

| Group | Participants in cohort | Person-months at risk (PMAR) | Reinfected – n (%) | Reinfection IR per 1,000 PMAR (95% CI) |
| --- | --- | --- | --- | --- |
| **All STH** |  |  |  |  |
| Pateque (improved WASH) | 112 | 534.82 | 44 (39.3%) | 82.27 (58.98-105.56) |
| Taninga (unimproved WASH) | 55 | 278.58 | 10 (18.2%) | 35.89 (14.05-57.74) |
| **Overall** | **167** | **813.40** | **54 (32.3%)** | **66.39 (49.29-83.49)** |
| ***T. trichiura*** |  |  |  |  |
| Pateque (improved WASH) | 70 | 329.24 | 24 (34.3%) | 72.89 (44.81-100.98) |
| Taninga (unimproved WASH) | 19 | 90.80 | 3 (15.8%) | 33.04 (-3.72-69.80) |
| **Overall** | **89** | **420.04** | **27 (30.3%)** | **64.28 (40.83-87.73)** |
| *A. lumbricoides* |  |  |  |  |
| Pateque (improved WASH) | 40 | 190.03 | 9 (22.5%) | 47.36 (17.16-77.56) |
| Taninga (unimproved WASH) | 17 | 79.03 | 6 (35.3%) | 75.92 (17.53-134.32) |
| **Overall** | **57** | **269.06** | **15 (26.3%)** | **55.75 (28.34-83.17)** |
| **Hookworms** |  |  |  |  |
| Pateque (improved WASH) | 15 | 79.97 | 5 (33.3%) | 62.52 (9.46-115.58) |
| Taninga (unimproved WASH) | 4 | 21.56 | 0 | 0 |
| **Overall** | **19** | **101.53** | **5 (26.3%)** | **49.24 (7.16-91.33)** |
| *S. stercoralis* |  |  |  |  |
| Pateque (improved WASH) | 3 | 8.57 | 2 (66.7%) | 233.40 (-49.82-516.63) |
| Taninga (unimproved WASH) | 22 | 116.02 | 1 (4.5%) | 25.44 (-8.20-25.44) |
| **Overall** | **25** | **124.59** | **3 (12%)** | **24.08 (-2.84-50.99)** |

787 IR – Incidence rate, PMAR – person-months at risk, CI – confidence interval.

Cox regression showed a statistically significant association between school and the hazard of reinfection for all STH combined (*p*=0.02). Children from the unimproved WASH cohort (Taninga) had a hazard ratio (HR) of 0.456 (95% CI: 0.229-0.908; *p*=0.0254) compared with those from the improved WASH cohort (Pateque). No statistically significant differences were observed between children enrolled in both schools for any specific STH species. Survival curves for all STH and individual species are shown in Figures 5-8 in Additional File 1.

## Discussion

We report the results of the WASH-it study, which aimed to (i) measure the efficacy of albendazole (with the addition of ivermectin when *T. trichiura* or *S. stercoralis* were present) against STH and (ii) describe STH reinfection rates over seven months following anthelmintic treatment, as a proxy for assessing the impact of a WASH improvement intervention. The study was conducted among school-aged children from two schools, following an intervention to improve WASH conditions in one of them. Overall, the baseline assessment revealed a higher prevalence of STH in Pateque (improved WASH) than in Taninga (unimproved WASH), with variation in the frequencies of STH species. After treatment, anthelmintic drugs showed high efficacy (≥80% CR and >90% ERR), regardless of STH species.

When the baseline assessment of this study was performed (first semester of 2019), mass drug administration of anthelmintics (albendazole and praziquantel) had been coordinated by the Ministry of Health and applied annually for approximately 8 years. In 2017, a cross-sectional study of the entire Manhiça district reported a prevalence of infection <20%, with less than 2% of infections classified as moderate to high intensity by Kato-Katz on a single stool sample (19). Here, we report even lower prevalences (all <10%) for each STH species, with no high-intensity infections detected. However, there were significant differences in baseline prevalence between the two schools where participants were enrolled. In contrast to previous reports (19), we observed different levels of infection intensity: for *T. trichiura* and hookworms, moderate-intensity infections accounted for 5-8% of the total, whereas for *A. lumbricoides*, they accounted for around 44% of infections. Baseline data from the ALIVE clinical trial (NCT 05124691) (26), conducted in Mozambique at the Pateque school between October 2022 and March 2023, were consistent with our observation. Overall, these findings indicate that, although the district as a whole was in 2017 closer to the WHO 2030 target of eliminating STH morbidity (indicated by infections of moderate and heavy intensity representing < 2% of infections) (7), the situation is more complex, with hotspots of moderate and heavy intensity infections in specific areas. This is also reflected in the distribution of STH species, with *T. trichiura* and *A. lumbricoides* significantly more frequent in Pateque, whereas the opposite was true for *S. stercoralis*. These findings underscore the need for a new, robust, and high-resolution national survey to assess the prevalence and intensity following MDA introduction and to evaluate its impact. This need is particularly timely, given that the last one was conducted two decades ago (27) and only a few locations have been surveyed since then (19,28–30). Such a survey is crucial to determine precisely how far Mozambique is from the WHO’ s 2030 STH elimination goals.

Our findings revealed that albendazole and ivermectin were effective against *T. trichiura*, with ERRs greater than 95%, well above the WHO-established reference efficacy of ≥50% for albendazole alone against *T. trichiura* (21). This finding, together with the CR of 88.4% we report for this parasite, indicates that this anthelmintic combination (albendazole and ivermectin) has the potential to significantly reduce the disease burden by decreasing infection intensities, as shown by recent trials (26,31–36). Similarly, for *A. lumbricoides*, the treatment efficacy was as high as reported in trials (all ERRs>99%), whether albendazole alone, a macrocyclic lactone (ivermectin or moxidectin) alone, or a combination of both or other drugs (26,32,37–39), indicating that albendazole alone remains effective against *A. lumbricoides*. Nonetheless, according to the WHO thresholds (21), the albendazole ERR, based on the arithmetic mean, is classified as doubtful (the ERR is lower than the reference efficacy by less than 10 percentage points). There are some reports of suspected reduced efficacy for A. lumbricoides in sub-Saharan Africa, particularly in Ethiopia and Rwanda (40,41); however, the data from our study should be interpreted with caution, given the relatively low number of participants infected with *A. lumbricoides* (*n*=60) assessed for treatment response. For hookworms and *S. stercoralis*, even though only a fraction of the recommended number of participants needed for treatment efficacy assessment (at least 50) were included, high cure rates and ERRs were observed for both, in line with other reports of efficacious treatment with albendazole for hookworms (26,32,38,40) and ivermectin for *S. stercoralis* (26,35), regardless of whether the drugs were administered separately or in combination.

Studies investigating STH reinfection rates after treatment with anthelmintic drugs have been conducted in multiple countries (42), including sub-Saharan Africa (36,40,43,44), but in Mozambique, most studies focused on pre-school-aged children (45). Here, the overall reinfection rate was 29.8% in school-aged children, with nearly one-third reinfected with STH seven months after anthelmintic treatment and cure. The reinfection rate in Pateque was more than double that of Taninga, even though the former had just received a WASH improvement intervention, including the provision of safe water, construction of improved latrines at the school, and community education and promotion of WASH. These reinfection rates might point to high levels of environmental contamination at the school and in the community of Pateque, but they may also be evidence that a school-based approach to deworming is not sufficient, because reservoirs of infection remain in the community (pre-school-aged children and adults not targeted by the MDA) (16). Another hypothesis is that, since WASH per se is not a driver of the reduction in the burden of STH, it will play a larger role in the long term in maintaining low morbidity by reducing reinfection rates, especially after interruption of preventive chemotherapy (12). Reinfection rates ranging from 37.2–52.4% (*T. trichiura*), 18.3–75.9% (*A. lumbricoides*) and 4.6–25% (hookworms) have been reported elsewhere in Africa (36,40,43), even though the study designs and time points for investigating reinfections varied (8 weeks to 1-year post-treatment). The reinfection rates reported in this study are approximately half of those estimated for around 6 months after infection had been cleared in a meta-analysis (46). The study design we employed enabled monthly detection and measurement of reinfection rates and estimation of reinfection incidence rates and hazard ratios. These analyses revealed that children from Taninga (school with unimproved WASH) were less frequently reinfected by STH than those enrolled in Pateque. This is more likely a reflection of differences in the epidemiological context across the two schools/regions of the district, raising the issue of differences in STH prevalence and infection intensity within the district, rather than necessarily reflecting the role WASH plays. Additionally, this study focused on school-aged participants included in the study, and did not take into consideration the range and frequency of WASH conditions an individual study participant is exposed to (e.g., at school vs. community), and much less the other members of their household (who may have professions that expose them to environments contaminated by STH), which suggests that WASH effectiveness depends on adequate coverage, sustained use, maintenance, behavioural uptake and reduction of environmental contamination beyond the school setting. Evidence from systematic reviews shows that WASH interventions may only modestly reduce STH infection, particularly when coverage is incomplete or when interventions are added to ongoing deworming programmes (14). Similarly, the WASH for WORMS trial in East Timor showed that adding a community-based WASH package to deworming did not produce clear additional reductions in STH infection, highlighting the importance of high coverage, long-term implementation and sustained open-defecation-free environments (15), as has also been postulated by a 7-year trial of WASH interventions that did not find consistent associations between WASH infrastructure and infection by STH (16).

Although progress has been made over the past decade, a large proportion of the population in Mozambique still has limited access to adequate water, sanitation, and hygiene (11). A previous study in Manhiça found that children under 15 years old in households with unimproved water and sanitation, but surrounded by neighbours with improved water and sanitation, had lower rates of outpatient visits for malaria, anaemia, and malnutrition (47). In the WASH-it study, we could not clearly demonstrate this association, largely due to the confounding factors mentioned above. It remains likely that improved WASH, in which sanitation reduces environmental contamination, hygiene reduces exposure to environmental infections, and water contributes to both, may be crucial, once elimination as a public health problem has been reached, to prevent recrudescence (12–14). These findings support integrated STH control packages rather than WASH as a stand-alone intervention. In areas with persistent reinfection, programmes may require fine-scale mapping, stronger surveillance, high treatment coverage, broader community-based deworming where appropriate, and more effective anthelmintic regimens. Recent evidence from the DeWorm3 trial supports the programmatic relevance of community-wide approaches for reducing STH transmission (48), while recent trials and the European Medicines Agency’s positive opinion on ivermectin/albendazole support the potential value of high-efficacy drug combinations for STH treatment, especially for *T. trichiura* and hookworm infections (26,37,49). The WHO also emphasises that WASH is a key component of integrated NTD control and should be planned, implemented, and evaluated jointly with disease-control programmes (50).

There are several limitations to our study. The main limitation is that we used a prospective cohort design comparing just two schools. This is compounded by heterogeneity in observed STH prevalence across the schools assessed, with a higher baseline prevalence in the school with improved WASH (Pateque) than in the school with unimproved WASH (Taninga). Improved WASH infrastructure (i.e., construction of clean and safe water and sanitation facilities) may not necessarily translate into improved practice and uptake (i.e., usage, maintenance, behaviour change). We did not measure community awareness and uptake of improved WASH, which could result in misleading exposure classification (based on the existence of infrastructure alone). Another consideration is other unmeasured factors (household and workplace WASH) that could further confound the WASH classification. These limitations shape any interpretations regarding the role of WASH in STH reinfection rates and reinfection incidence rates. Due to differences in baseline STH prevalence, it is difficult to compare the two schools’ reinfection rates directly. Due to the lower prevalence, the small sample size was insufficient to precisely measure efficacy and treatment response for hookworms and *S. stercoralis*. Finally, reliance on a single microscopy-based stool sample may have reduced sensitivity for detecting low-intensity infections, particularly in post-treatment and reinfection assessments. This may have resulted in missed residual or newly acquired infections, thereby overestimating treatment efficacy while underestimating reinfections.

## Conclusion

Our findings highlight variations in STH prevalence, infection burden, and reinfection rates across the Manhiça district. Although the treatment response assessment indicated good efficacy, reinfection occurred within seven months of treatment in more than a third of participants, even in an area with improved WASH conditions and community-based education campaigns. These findings underscore the need for robust, fine-scale surveys to monitor the impact of MDA, supported by cluster-randomised trials or quasi-experimental studies (51) to investigate the epidemiological and programmatic implications of STH reinfection and the role of WASH in interrupting transmission.

## Declarations

### Ethics approval and consent to participate

The WASH-it study protocol, data collection tools, and informed consent were reviewed and approved by CISM’s Institutional Bioethics Committee – CIBS-CISM (002/2018) and by the Mozambican National Bioethics Committee for Health – CNBS (Ref: 199/CNBS/18). After informing each child’s caretaker of the objectives and characteristics of the study, two copies of a written informed consent form were signed by those who agreed to participate (or by an impartial witness when the parent/guardian was illiterate) after an information sheet was provided and read to them. Ample time was given to present doubts or questions, which the CISM investigator clarified. One copy was given to the caretaker, and the other was stored in a secure archive at the Centro de Investigação em Saúde de Manhiça (CISM). Children aged 12 to 17 were also asked to sign an informed assent form. Participation in the WASH-it study was voluntary, and the guardians were free to withdraw the child from the study at any time. The study was conducted in accordance with the ICH GCP guidelines. All children found to have treatable conditions were referred to a local health facility (or the Manhiça District Hospital) and received treatment according to national treatment protocols. Clinical trial number: not applicable.

### Consent for publication

Not applicable.

### Availability of data and materials

The datasets for the current study are available from the corresponding author on reasonable request, accompanied by a short formal proposal addressed to, that will be analysed by CISM’s internal scientific and ethical committees.

### Authors’ contributions

Conceptualization: VN, BGP, CS, JM; Data curation: VN, AMJ, AC, OM; Formal analysis: AMJ, PEF, SRD; Funding acquisition: JM; Investigation: VN, AMJ, BGP, JG, CS, AC, OM, JCJ; Methodology: VN, AMJ, BGP, CS, OM, JM; Project administration: VN, BGP, OM, JM; Resources: VN, AC, OM, KM, IM, JM; Software: AMJ, PEF, SRD; Supervision: BGP, JG, MMV, KM, IM, SRD, JM; Validation: PEF, JG, MMV; Visualization: AMJ, PEF, SRD, JM; Writing – original draft preparation: VN, AMJ, PEF, SRD, JM; Writing – review and editing: VN, AMJ, PEF, BGP, JG, CS, AC, OM, JCJ, MMV, KM, IM, SRD, JM. All authors contributed to the article and approved the submitted version. All authors have read and agreed to the published version of the manuscript.

## List of abbreviations

ALB: Albendazole
CI: Confidence interval
CISM: Centro de Investigação em Saúde de Manhiça
CR: Cure rate
EPG: Eggs per gram of stool
ERR: Egg reduction rate
HDSS: Health and Demographic Surveillance System
HR: Hazard ratio
IR: Incidence rate
IVM: Ivermectin
KK: Kato-Katz
MDA: Mass Drug Administration
NGO: Non-Government Organization
NTD: Neglected Tropical Diseases
PMAR: Person-months at risk
Pre-SAC: Pre-school aged children
PZQ: Praziquantel
SAC: School-aged children
STH: Soil-transmitted helminth
UNICEF: United Nations Children’s Fund
WASH: Water, Sanitation, and Hygiene
WHO: World Health Organization

## Acknowledgements

We thank all WASH-it study participants, as well as the pupils, school staff, and community members who generously participated in this research. We are also grateful to the district authorities and to our colleagues at the Centro de Investigação em Saúde de Manhiça (CISM) for their continuous support and to the NGO ONGAWA, which collaborated with us in this study. CISM receives core funding from the Spanish Agency for International Cooperation – AECID (Ministry of Foreign Affairs, Spain). ISGlobal authors acknowledge support from the grant CEX2023-0001290-S funded by the MCIN/AEI/10.13039/501100011033, and support from the Generalitat de Catalunya through the CERCA Program. SRD and JG are supported by a UKRI Future Leaders Fellowship [grant: MR/T020733/1 to SRD], the Wellcome Trust (UK) through core funding to the Wellcome Sanger Institute (UK) [grant: 220540/Z/20/A], and STOP2030, an EDCTP3 programme [grant: 101103089] supported by the European Union.

## Authors’ information

VN and AMJ contributed equally to this work and are joint first authors.

## References

1. Jourdan PM, Lamberton PHL, Fenwick A, Addiss DG. Soil-transmitted helminth infections. The Lancet. 2018 Jan;391(10117):252–65. doi:10.1016/S0140-6736(17)31930-X PubMed PMID: 28882382.

2. Hotez PJ. “The Unholy Trinity”: the Soil-Transmitted Helminth Infections Ascariasis, Trichuriasis, and Hookworm Infection. In: Hotez PJ, editor. Forgotten People, Forgotten Diseases: The Neglected Tropical Diseases and Their Impact on Global Health and Development. 3rd ed. Washington DC, USA: American Society for Microbiology Press and John Wiley & Sons, Inc.; 2021. p. 17–38. doi:10.1128/9781683673903.ch02

3. Chen J, Gong Y, Chen Q, Li S, Zhou Y. Global burden of soil-transmitted helminth infections, 1990-2021. Infect Dis Poverty. 2024 Oct 24;13(1):77. doi:10.1186/s40249-024-01238-9 PubMed PMID: 39444032.

4. Krolewiecki A, Nutman TB. Strongyloidiasis: A Neglected Tropical Disease. Infect Dis Clin North Am. 2019 Mar 1;33(1):135–51. doi:10.1016/j.idc.2018.10.006 PubMed PMID: 30712758.

5. Fleitas PE, Travacio M, Martí-Soler H, Socías ME, Lopez WR, Krolewiecki AJ. The strongyloides stercoralis-hookworms association as a path to the estimation of the global burden of strongyloidiasis: A systematic review. PLoS Negl Trop Dis. 2020 Apr 1;14(4):1–13. doi:10.1371/journal.pntd.0008184 PubMed PMID: 32282827.

6. Crompton DWT, Nesheim MC. Nutritional impact of intestinal helminthiasis during the human life cycle. Annual Review of Nutrition. 2002. p. 35–59. doi:10.1146/annurev.nutr.22.120501.134539 PubMed PMID: 12055337.

7. World Health Organization. 2030 Targets for Soil-Transmitted Helminthiases Control Programmes. World Health Organization; 2020.

8. World Health Organization. Guideline: preventive chemotherapy to control soil-transmitted helminth infections in at-risk population groups. 2017.

9. WHO Control of Neglected Tropical Diseases Team. Ending the Neglect to Attain the Sustainable Development Goals: A Road Map for Neglected Tropical Diseases 2021-2030. World Health Organization; 2020. 196 p.

10. Keenan JD, Hotez PJ, Amza A, Stoller NE, Gaynor BD, Porco TC, et al. Elimination and Eradication of Neglected Tropical Diseases with Mass Drug Administrations: A Survey of Experts. PLoS Negl Trop Dis. 2013 Dec 5;7(12):e2562. doi:10.1371/journal.pntd.0002562

11. WHO/UNICEF Joint Monitoring Programme Team. Progress on household drinking water, sanitation and hygiene 2000-2025: special focus on inequalities [Internet]. Geneva; 2025 Aug [cited 2026 Jan 8]. Available from: https://www.who.int/publications/m/item/progress-on-household-drinking-water--sanitation-and-hygiene-2000-2024--special-focus-on-inequalities

12. Coffeng LE, Vaz Nery S, Gray DJ, Bakker R, de Vlas SJ, Clements ACA. Predicted short and long-term impact of deworming and water, hygiene, and sanitation on transmission of soil-transmitted helminths. PLoS Negl Trop Dis. 2018 Dec 1;12(12). doi:10.1371/journal.pntd.0006758 PubMed PMID: 30522129.

13. Strunz EC, Addiss DG, Stocks ME, Ogden S, Utzinger J, Freeman MC. Water, Sanitation, Hygiene, and Soil-Transmitted Helminth Infection: A Systematic Review and Meta-Analysis. PLoS Med. 2014;11(3). doi:10.1371/journal.pmed.1001620 PubMed PMID: 24667810.

14. Garn J V., Wilkers JL, Meehan AA, Pfadenhauer LM, Burns J, Imtiaz R, et al. Interventions to improve water, sanitation, and hygiene for preventing soil-transmitted helminth infection. Cochrane Database of Systematic Reviews. John Wiley and Sons Ltd; 2022. doi:10.1002/14651858.CD012199.pub2 PubMed PMID: 35726112.

15. Vaz Nery S, Traub RJ, McCarthy JS, Clarke NE, Amaral S, Llewellyn S, et al. WASH for WORMS: A Cluster-Randomized Controlled Trial of the Impact of a Community Integrated Water, Sanitation, and Hygiene and Deworming Intervention on Soil-Transmitted Helminth Infections. Am J Trop Med Hyg. 2019 Mar 6;100(3):750–61. doi:10.4269/ajtmh.18-0705

16. Maddren R, Liyew EF, Anjulo U, Alemu ZA, Chernet M, Collyer BS, et al. Measuring the impact of water, sanitation, and hygiene (WaSH) infrastructure upon soil-transmitted helminth infection in southern Ethiopia: individual-level analysis of the Geshiyaro project after 7 years. The Lancet Regional Health – Africa. 2026 May;3:100049. doi:10.1016/j.lanafr.2026.100049

17. Sacoor C, Nhacolo A, Nhalungo D, Aponte JJ, Bassat Q, Sacarlal J, et al. Profile: Manhiça Health Research Centre (Manhiça HDSS). Int J Epidemiol. 2013;42(5):1309–18. doi:10.1093/ije/dyt148 PubMed PMID: 24159076.

18. Nhacolo A, Jamisse E, Augusto O, Matsena T, Hunguana A, Mandomando I, et al. Cohort profile update: Manhiça health and demographic surveillance system (HDSS) of the Manhiça health research centre (CISM). Int J Epidemiol. 2021 Jan 16;1–8. doi:10.1093/ije/dyaa218

19. Grau-Pujol B, Martí-Soler H, Escola V, Demontis M, Jamine JC, Gandasegui J, et al. Towards soil-transmitted helminths transmission interruption: The impact of diagnostic tools on infection prediction in a low intensity setting in Southern Mozambique. Beechler BR, editor. PLoS Negl Trop Dis. 2021 Oct 25;15(10):e0009803. doi:10.1371/journal.pntd.0009803 PubMed PMID: 34695108.

20. Kar Kamal, Chambers Robert. Handbook on community-led total sanitation. London: Plan UK & Institute of Development Studies; 2008. 96 p.

21. WHO Control of Neglected Tropical Diseases Team. Assessing the efficacy of anthelminthic drugs against schistosomiasis and soil-transmitted helminthiases. Montresor A, editor. Geneva, Switzerland: World Health Organization; 2013. 1–39 p.

22. Aramendia AA, Anegagrie M, Zewdie D, Dacal E, Saugar JM, Herrador Z, et al. Epidemiology of intestinal helminthiases in a rural community of Ethiopia: Is it time to expand control programs to include Strongyloides stercoralis and the entire community? PLoS Negl Trop Dis. 2020 Jun 4;14(6):e0008315. doi:10.1371/journal.pntd.0008315

23. WHO Expert Committee on the Control of Schistosomiaiss. Prevention and control of schistosomiasis and soil-transmitted helminthiasis: report of a WHO expert committee. Geneva, Switzerland: World Health Organization; 2002. 57 p.

24. Sacko M, De Clercq D, Behnke JM, Gilbert FS, Dorny P, Vercruysse J. Comparison of the efficacy of mebendazole, albendazole and pyrantel in treatment of human hookworm infections in the Southern Region of Mali, West Africa. Trans R Soc Trop Med Hyg. 1999 Mar;93(2):195–203. doi:10.1016/S0035-9203(99)90306-1

25. R Core Team. R: A Language and Environment for Statistical Computing. Vienna, Austria: R Foundation for Statistical Computing; 2025.

26. Krolewiecki A, Kepha S, Fleitas PE, van Lieshout L, Gelaye W, Messa Jr. A, et al. Albendazole– ivermectin co-formulation for the treatment of Trichuris trichiura and other soil-transmitted helminths: a randomised phase 2/3 trial. Lancet Infect Dis. 2025 May;25(5):548–59. doi:10.1016/S1473-3099(24)00669-8 PubMed PMID: 39805305.

27. Augusto G, Nalá R, Casmo V, Sabonete A, Mapaco L, Monteiro J. Geographic distribution and prevalence of schistosomiasis and soil-transmitted helminths among schoolchildren in mozambique. American Journal of Tropical Medicine and Hygiene. 2009;81(5):799–803. doi:10.4269/ajtmh.2009.08-0344 PubMed PMID: 19861614.

28. Meurs L, Polderman AM, Vinkeles Melchers NVS, Brienen EAT, Verweij JJ, Groosjohan B, et al. Diagnosing Polyparasitism in a High-Prevalence Setting in Beira, Mozambique: Detection of Intestinal Parasites in Fecal Samples by Microscopy and Real-Time PCR. PLoS Negl Trop Dis. 2017;11(1):1–18. doi:10.1371/journal.pntd.0005310 PubMed PMID: 28114314.

29. Alfredo C, Nchowela GA, Mabasso AA, Muchanga IJ, Muadica AS. Prevalence of Schistosoma haematobium and Soil-Transmitted Helminths Infections among School-Aged Children in Quelimane and Gurue Districts, Central Mozambique. J Bacteriol Prasitol. 2022;13:22. doi:10.35248/2155-9597.22.S18.022

30. Casmo V, Chicumbe S, Chambisse R, Nalá R. Regional Differences in Intestinal Parasitic Infections among Army Recruits in a Southern Mozambique Training Center: A Cross-Sectional Study. Pathogens. 2023 Sep 1;12(9). doi:10.3390/pathogens12091105

31. Matamoros G, Sánchez A, Gabrie JA, Juárez M, Ceballos L, Escalada A, et al. Efficacy and Safety of Albendazole and High-Dose Ivermectin Coadministration in School-Aged Children Infected With *Trichuris trichiura* in Honduras: A Randomized Controlled Trial. Clinical Infectious Diseases. 2021 Oct 5;73(7):1203–10. doi:10.1093/cid/ciab365

32. Knopp S, Mohammed KA, Speich B, Hattendorf J, Khamis IS, Khamis AN, et al. Albendazole and Mebendazole Administered Alone or in Combination with Ivermectin against *Trichuris trichiura*: A Randomized Controlled Trial. Clinical Infectious Diseases. 2010 Dec 15;51(12):1420–8. doi:10.1086/657310

33. Palmeirim MS, Hürlimann E, Knopp S, Speich B, Belizario V, Joseph SA, et al. Efficacy and safety of co-administered ivermectin plus albendazole for treating soil-transmitted helminths: A systematic review meta-analysis and individual patient data analysis. PLoS Negl Trop Dis. 2018;12(4):1–26. doi:10.1371/journal.pntd.0006458

34. Palmeirim MS, Hürlimann E, Beinamaryo P, Kyarisiima H, Nabatte B, Hattendorf J, et al. Efficacy and safety of albendazole alone versus albendazole in combination with ivermectin for the treatment of Trichuris trichiura infections: An open-label, randomized controlled superiority trial in south-western Uganda. Deye G, editor. PLoS Negl Trop Dis. 2024 Nov 26;18(11):e0012687. doi:10.1371/journal.pntd.0012687

35. Speich B, Ali SM, Ame SM, Bogoch II, Alles R, Huwyler J, et al. Efficacy and safety of albendazole plus ivermectin, albendazole plus mebendazole, albendazole plus oxantel pamoate, and mebendazole alone against Trichuris trichiura and concomitant soil-transmitted helminth infections: a four-arm, randomised controlled trial. Lancet Infect Dis. 2015 Mar;15(3):277–84. doi:10.1016/S1473-3099(14)71050-3

36. Speich B, Moser W, Ali SM, Ame SM, Albonico M, Hattendorf J, et al. Efficacy and reinfection with soil-transmitted helminths 18-weeks post-treatment with albendazole-ivermectin, albendazole-mebendazole, albendazole-oxantel pamoate and mebendazole. Parasit Vectors. 2016 Mar 2;9(1). doi:10.1186/s13071-016-1406-8 PubMed PMID: 26935065.

37. Hürlimann E, Hofmann D, Keiser J. Ivermectin and moxidectin against soil-transmitted helminth infections. Trends Parasitol. 2023 Apr 1;39(4):272–84. doi:10.1016/j.pt.2023.01.009

38. Hürlimann E, Keller L, Patel C, Welsche S, Hattendorf J, Ali SM, et al. Efficacy and safety of co-administered ivermectin and albendazole in school-aged children and adults infected with Trichuris trichiura in Côte d’Ivoire, Laos, and Pemba Island, Tanzania: a double-blind, parallel-group, phase 3, randomised controlled trial. Lancet Infect Dis. 2022 Jan;22(1):123–35. doi:10.1016/S1473-3099(21)00421-7

39. Speich B, Ali SM, Ame SM, Albonico M, Utzinger J, Keiser J. Quality control in the diagnosis of Trichuris trichiura and Ascaris lumbricoides using the Kato-Katz technique: Experience from three randomised controlled trials. Parasit Vectors. 2015;8(1):1–8. doi:10.1186/s13071-015-0702-z

40. Gebreyesus TD, Makonnen E, Tadele T, Mekete K, Gashaw H, Gerba H, et al. Reduced efficacy of single-dose albendazole against Ascaris lumbricoides, and Trichuris trichiura, and high reinfection rate after cure among school children in southern Ethiopia: a prospective cohort study. Infect Dis Poverty. 2024 Dec 1;13(1). doi:10.1186/s40249-024-01176-6 PubMed PMID: 38246985.

41. Krücken J, Fraundorfer K, Mugisha JC, Ramünke S, Sifft KC, Geus D, et al. Reduced efficacy of albendazole against Ascaris lumbricoides in Rwandan schoolchildren. Int J Parasitol Drugs Drug Resist. 2017 Dec 1;7(3):262–71. doi:10.1016/j.ijpddr.2017.06.001 PubMed PMID: 28697451.

42. Yap P, Utzinger J, Hattendorf J, Steinmann P. Influence of nutrition on infection and re-infection with soil-transmitted helminths: A systematic review. Parasites and Vectors. BioMed Central Ltd.; 2014. doi:10.1186/1756-3305-7-229 PubMed PMID: 24885622.

43. Zeleke AJ, Bayih AG, Afework S, Gilleard JS. Treatment efficacy and re-infection rates of soil-transmitted helminths following mebendazole treatment in schoolchildren, Northwest Ethiopia. Trop Med Health. 2020 Dec 1;48(1). doi:10.1186/s41182-020-00282-z

44. Gomez SR, Maddren R, Liyew EF, Chernet M, Anjulo U, Tamiru A, et al. Predisposition to soil-Transmitted helminth reinfection after four rounds of mass drug administration: Results from a longitudinal cohort in the Geshiyaro project, a transmission elimination feasibility study in the Wolaita zone of southern Ethiopia. Trans R Soc Trop Med Hyg. 2023 Jul 1;117(7):514–21. doi:10.1093/trstmh/trad007 PubMed PMID: 36939014.

45. Knee J, Sumner T, Adriano Z, Anderson C, Bush F, Capone D, et al. Effects of an urban sanitation intervention on childhood enteric infection and diarrhea in maputo, mozambique: A controlled before-and-after trial. Elife. 2021 Apr 1;10. doi:10.7554/ELIFE.62278 PubMed PMID: 33835026.

46. Jia TW, Melville S, Utzinger J, King CH, Zhou XN. Soil-Transmitted Helminth Reinfection after Drug Treatment: A Systematic Review and Meta-Analysis. Cooper PJ, editor. PLoS Negl Trop Dis. 2012 May 8;6(5):e1621. doi:10.1371/journal.pntd.0001621

47. Grau-Pujol B, Cano J, Marti-Soler H, Casellas A, Giorgi E, Nhacolo A, et al. Neighbors’ use of water and sanitation facilities can affect children’s health: a cohort study in Mozambique using a spatial approach. BMC Public Health. 2022 Dec 1;22(1). doi:10.1186/s12889-022-13373-9 PubMed PMID: 35578273.

48. Ajjampur SSR, Aruldas K, Ásbjörnsdóttir KH, Avokpaho E, Bailey R, Cottrell G, et al. Feasibility of interrupting the transmission of soil-transmitted helminths: the DeWorm3 community cluster-randomised controlled trial in Benin, India, and Malawi. The Lancet. 2025 Aug 2;406(10502):475–88. doi:10.1016/S0140-6736(25)00766-4

49. European Medicines Agency. New combination of medicines to treat parasitic worm infections [Internet]. 2025 [cited 2026 Jun 8]. Available from: https://www.ema.europa.eu/en/news/new-combination-medicines-treat-parasitic-worm-infections

50. World Health Organization. Ending the Neglect to Attain the Sustainable Development Goals: A Global Strategy on Water, Sanitation and Hygiene to Combat Neglected Tropical Diseases, 2021-2030. World Health Organization; 2021.

51. Bärnighausen T, Oldenburg C, Tugwell P, Bommer C, Ebert C, Barreto M, et al. Quasi-experimental study designs series—paper 7: assessing the assumptions. J Clin Epidemiol. 2017 Sep;89:53–66. doi:10.1016/j.jclinepi.2017.02.017

